# Differential Effects of Cognitive vs. Motor Dual-Task Training in Stroke Rehabilitation: A Precision-Focused Meta-Analysis

**DOI:** 10.64898/2026.01.23.26344517

**Authors:** Hui Gao, Man Lang, Mustapha Mangdow, Wen Liu

**Author notes:** Co-first authors (Hui Gao, Man Lang).

## Abstract

This systematic review and meta-analysis primarily aimed to investigate the differential effectiveness of motor dual-task training (MDT) and cognitive dual-task training (CDT) on gait performance, balance control, and motor function in stroke survivors, and explored other important moderating factors such as stroke chronicity and individual functional profiles to inform a precision-based, personalized approach. Twenty-one RCTs involving 786 stroke survivors were included. Dual-task training demonstrated a medium overall beneficial effect on both temporal and spatial gait performance (SMD=0.50, p=0.03; SMD=0.5, p=0.04) and balance control (SMD=0.71, p=0.02), whereas no statistically significant improvement was observed in lower-extremity motor function. Subgroup analysis revealed that dual-task training modality was a critical determinant of treatment response. MDT was significantly superior for gait performance on both gait speed and stride length (SMD=1.15, p=0.01; SMD=0.89, p<0.01), while CDT demonstrated a significant benefit for balance control (SMD=0.59, p<0.01). Those modality-specific effects were further supported by meta-regression analysis. Stroke survivors at high risk of falls showed greater balance improvements following dual-task training. Furthermore, improvements in balance control and motor function were observed in non-chronic stroke survivors (≤6 months post-stroke) but not in chronic stroke survivors. These results offer crucial prescriptive insights, guiding clinicians to match the dual-task modality and timing of intervention to the individual patient’s functional profile. However, the high heterogeneity among studies and the lack of direct comparative trials between CDT and MDT limit the conclusive strength of these recommendations.

## Introduction

Stroke is one of the leading causes of disability^1^. Most stroke survivors show impairment in gait pattern, trunk posture, and balance control, as well as cognitive and perceptual function^2^. Balance control is the ability to maintain body movement within the base of support without falling, which can be influenced by cognitive factors, such as attention, motivation, and intent^3^. Gait impairment consists of slow walking speed and short duration, and affects activities of daily living (ADL) in stroke survivors^4^. Most of the ADLs require the ability to perform two or more tasks simultaneously^5^. Therefore, dual-task training involving performing two or more tasks at the same time has been widely applied and evaluated in stroke rehabilitation^6^.

However, the evidence supporting the benefits of dual-task training in improving overall physical function after stroke remains mixed. This inconsistency may be due to the way that dual-task training is often treated as a single, uniform intervention, despite wide variation in both patient characteristics and training methods. A meta-analysis^7^ conducted in 2021, indicated significant benefits of dual-task training on balance, gait, and upper limb function compared with conventional therapy, whereas a 2022 meta-analysis^6^ reported no significant difference on balance outcome between the comparative groups. Dual-task studies are often classified into two different types: cognitive dual tasks (CDT) or motor dual tasks (MDT), which respectively refer to the simultaneous performance of one cognitive task and one motor task^8^, or simultaneous performance of two different motor tasks^9^. Only two past studies directly compared two different types of dual-task training. Inconsistent findings were reported either better outcomes after the CDT program than the MDT program^10^, or the opposite^11^. Furthermore, patient characteristics such as stroke chronicity and baseline function, both of which are known to influence treatment response, were not accounted for in prior meta-analyses.

Aligned with the NIH milestones for advancing precision medicine in rehabilitation^12^, this systematic review and meta-analysis is structured around three objectives. The primary objective is to examine the differential effectiveness of two dual-task training modalities, CDT and MDT, on gait, balance control, and motor function in stroke survivors. Given the heterogeneity of post-stroke functional impairments, clarifying these differential effects may facilitate personalized prescription of dual-task training tailored to specific functional deficits, thereby moving beyond a “one-size-fits-all” rehabilitation approach. The secondly objective is to explore potential moderators of training effects, including participant characteristics and types of control interventions. Stroke survivors with different levels of chronicity or baseline functional impairment severity in walking or balance control, may respond differently to dual-task training.

Investigating these moderating factors can further support the implementation of dual-task interventions within a precision medicine framework. Finally, by including several newly published clinical trial reports^13–17^ of dual-task training, this review and meta-analysis study provides an updated quantitative synthesis of the overall effects of dual-task training in stroke rehabilitation.

## Methods

This study was registered in the International Prospective Register of Systematic Reviews (PROSPERO) (Registration ID: CRD42023477417).

This review was performed and reported according to the Preferred Reporting Items for Systematic Reviews and Meta-Analysis (PRISMA) statement^18^. Two investigators independently performed the literature search via six English databases (Web of Science, PubMed, Medline, Embase, Cochrane Library, CINAHL) and one Chinese database (Chinese National Knowledge Infrastructure). The search terms were stroke/CVA/cerebrovascular accident and dual-task. The literature search commenced in November 2023 and concluded in June 2025 The inclusion criteria for study selection were (1) population of stroke survivors; (2) randomized controlled trials that used dual-task training compared with conventional therapy or sham control; (3) outcomes including one or more of the following: temporal gait parameter (gait speed), spatial gait parameter (stride length), balance (Berg Balance Scale (BBS)), and motor function (Fugl-Meyer Lower Extremity Assessment score (FMA_LA)); (4) full-text studies published in English, or Chinese articles with English abstract.

Exclusion criteria were (1) healthy population or non-stroke patients as comparative participants; (2) dual-task training combining with other confounding training such as aquatic exercise; (3) protocols, reviews, letters, academic thesies, and congress abstracts; (4) studies that outcome data (e.g., mean ± SD) could not be extracted from the text, tables, or figures, and author contact was unsuccessful.

After two researchers independently screened literature and identified a final list of the qualified studies based on inclusion and exclusion criteria, data extraction and statistical analysis were performed according to the Cochrane Handbook for Systematic Reviews of Interventions^19^. The following data were extracted: sample size, patients’ characteristics, intervention characteristics, and outcomes. To facilitate the subgroup analyses, patient characteristics such as fall risk and walking ability were defined using baseline scores from the BBS, and gait speed, respectively. Fall risk was categorized based on BBS scores as follows: low risk (41–56), moderate risk (21–40), and high risk(0-20)^20^. Walking ability was assessed using comfortable gait speed, with 0.49 m/s serving as the threshold to distinguish between home ambulators and community ambulators^22^. In terms of the intervention characteristics, dual-task training was categorized into CDT and MDT. The type of basic motor training was classified as gait training, balance training, or mixed training. For instance, if the basic motor component of dual-task training involved walking, the intervention was categorized as dual-task gait training. The types of control were classified as either active or passive. Active control means comparable standard treatment, and passive control refers to no additional intervention other than usual care.

The methodological quality of studies was appraised by two reviewers independently using the Physiotherapy Evidence Database (PEDro) scale. Studies with a score of six or above were classified as high quality, whereas scores of four and five were classified as fair quality, and scores below three were considered poor quality^23^. Discrepancies that could not be resolved through discussion were evaluated by a third expert.

StataSE15 was used to conduct meta- and subgroup analysis on three outcomes: gait performance (gait speed and stride length), balance (BBS), and motor function (FMA_LE). After completing the analysis, relevant values for each parameter, such as p-values, standardized mean differences (SMD), etc., were entered into Microsoft Excel to create a visually appealing forest plot that summarizes the descriptive data^24^.

Heterogeneity was assessed using the I² statistic, with the following interpretation: 0–40% low heterogeneity, 30–60% moderate heterogeneity, 50–90% substantial heterogeneity, and 75–100% considerable heterogeneity^19^. When an I² value is greater than 50%, a random-effects model would be applied; otherwise, a fixed-effects model would be used. Pooled effect sizes were calculated with z tests, where a p-value <0.05 demonstrated statistical significance. We used Hedges’ g with 95% confidence interval (CI) to calculate effect sizes, as it corrects for small sample sizes, which were categorized as follows: small (≥ 0.2), medium (≥ 0.5), and large (≥ 0.8)^25^.

Comprehensive subgroup analyses were conducted to examine the differential effectiveness of the dual-task training. Key comparisons included the training modalities (CDT vs. MDT), stroke chronicity (< 6 months vs. ≥ 6 months), fall risk levels (low vs. moderate vs. high), walking ability (home ambulator vs. community ambulator), type of basic motor training (gait training vs. balance training vs. mixed training), and type of control method (active vs. passive control).To ensure statistical robustness, any subgroup containing only a single comparison or insufficient data to calculate heterogeneity or p-values was excluded. Furthermore, we conducted meta-regression on the dual-task modalities (CDT vs. MDT) to statistically justify the differential findings observed in the subgroup analysis (results are presented in the Supplementary Materials).

Sensitivity analyses were performed to assess the stability of the primary findings and identify studies disproportionately contributing to heterogeneity. To assess potential publication bias, Egger’s regression test was conducted. It is a quantitative test with p< 0.05 indicating significant publication bias.

## Results

Figure 1 shows the process of literature search. An initial total of 1405 articles were identified. After removing duplications, systematic reviews, and meta-analyses, 1195 articles were screened for eligibility, and 21 articles were included for the final review. Of those studies, three^26–28^ were in Chinese and 18^13–17,29–41^ were in English.

**Figure 1.**
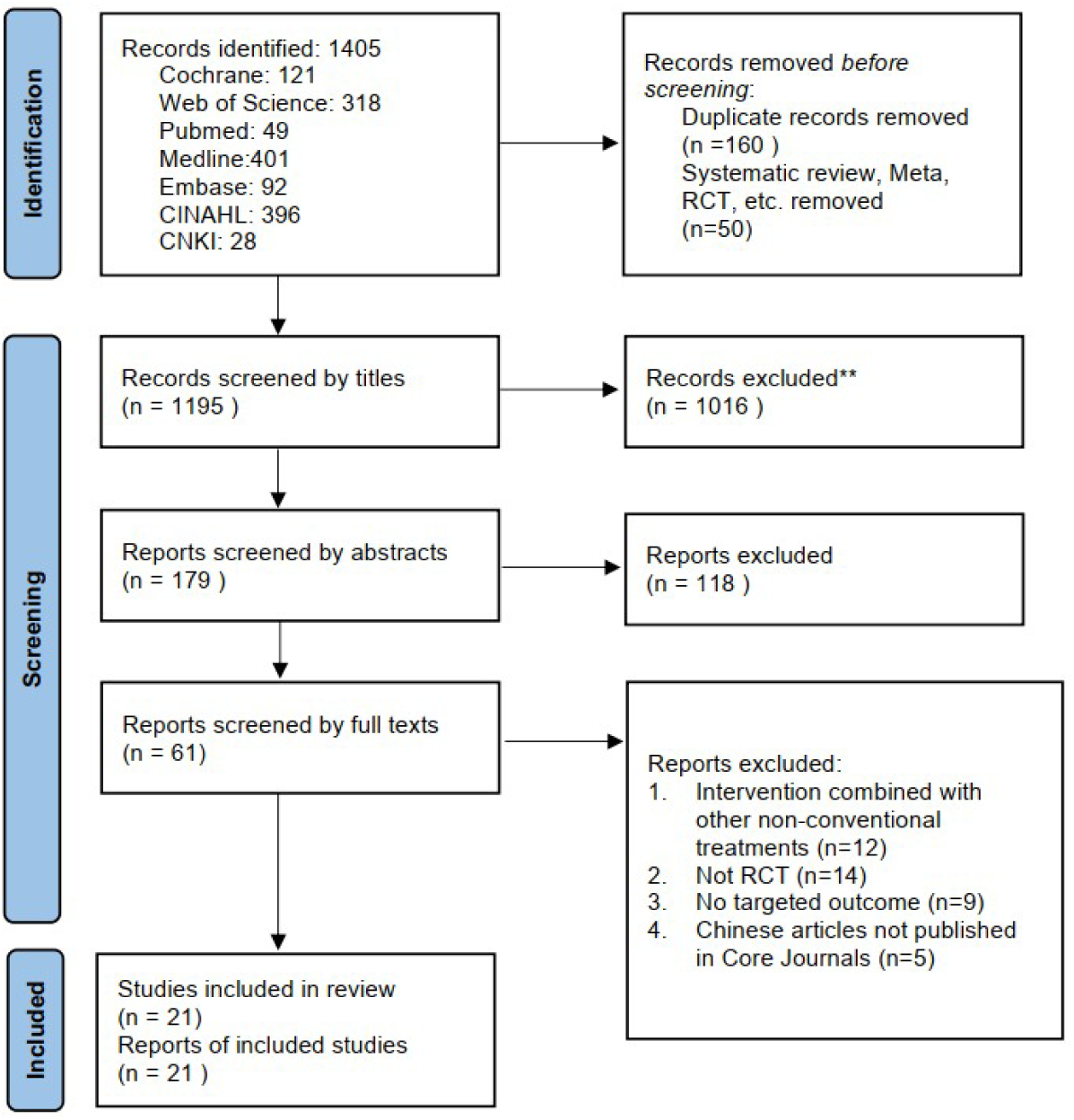
PRISMA flow diagram.

These studies were published between 2007 and 2025, and the study locations included the USA, Pakistan, Turkey, South Korea, Taiwan, and China. A total of 786 participants were included in this review, with 396 participants allocated to the dual-task group and 390 to the control group. None of the included studies reported significant baseline differences between comparative groups in terms of gender distribution, age, or disease duration. In most studies, there were no significant baseline differences in outcome measures between control and experiment groups. In only one study^33^, the baseline stride length of the control group was almost twice that of the experiment group. The detailed characteristics of participants and intervention of the included studies are summarized in Table 1 and Table 2, respectively.

**Table 1.**
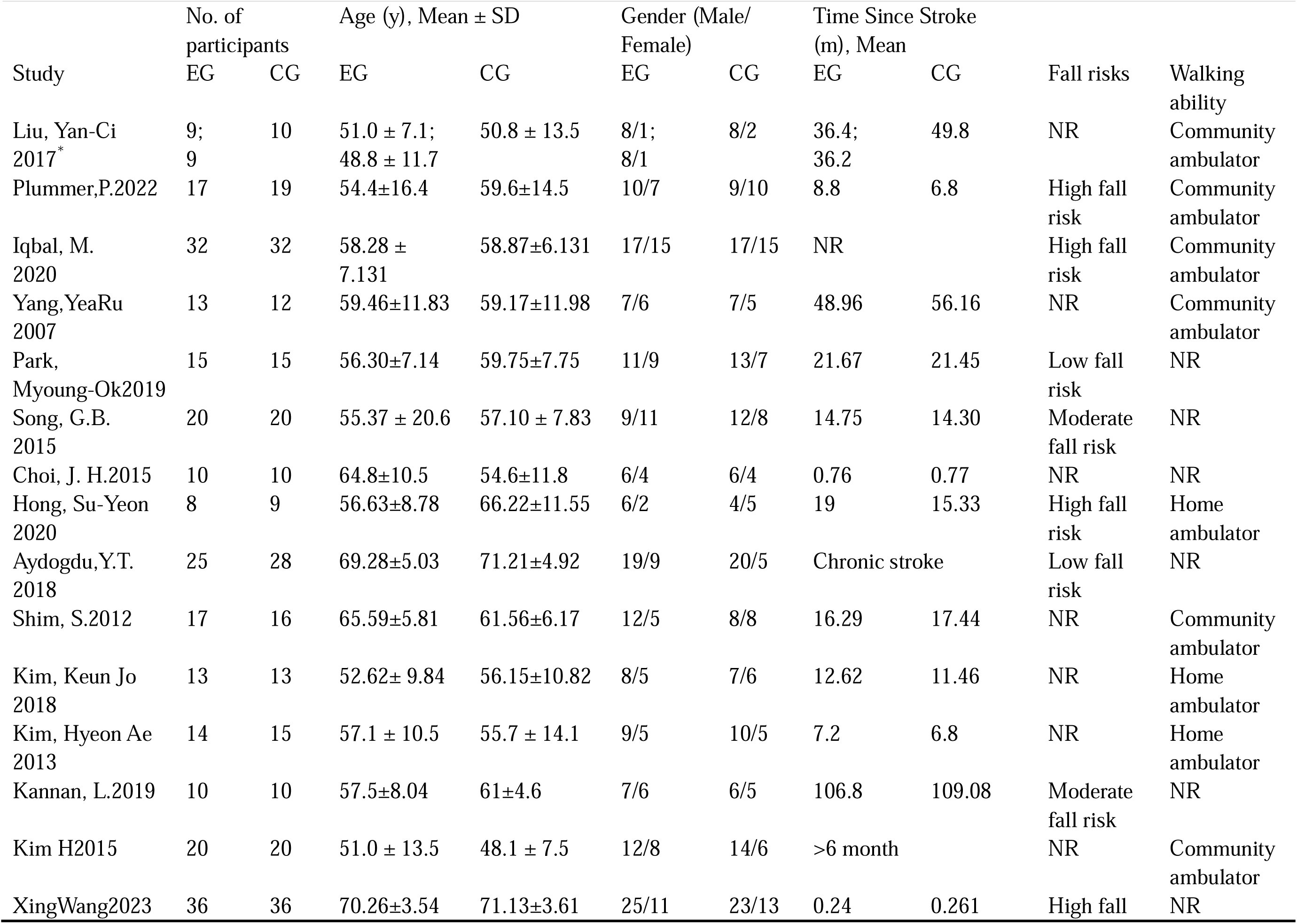
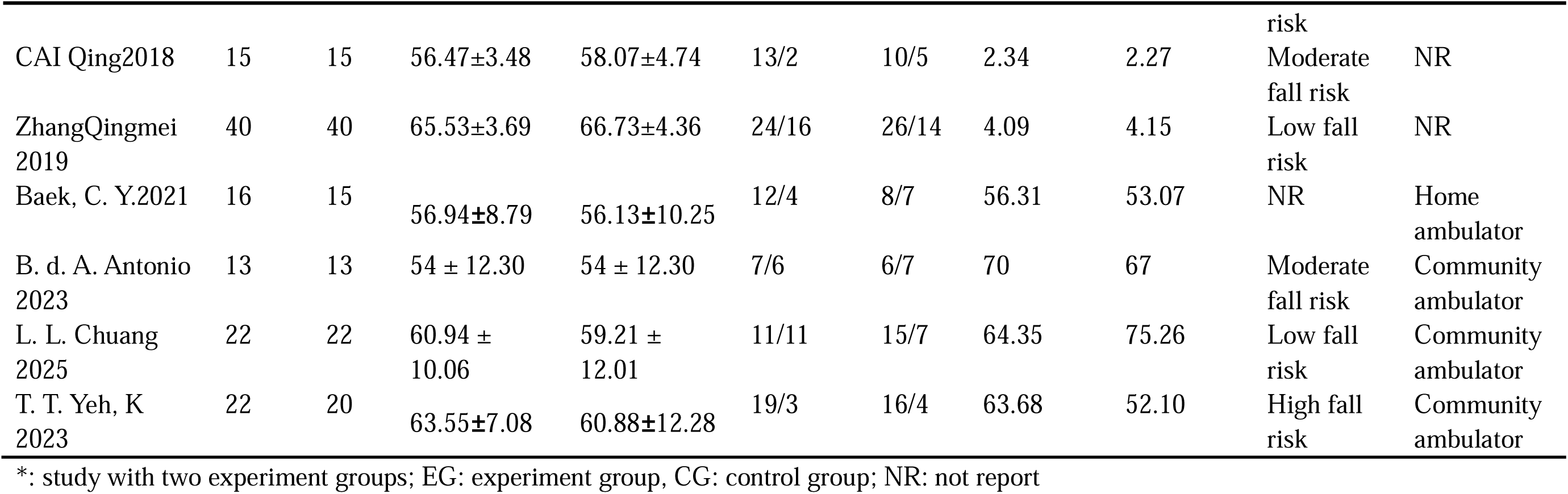
Participant characteristics of the included studies

**Table 2.**
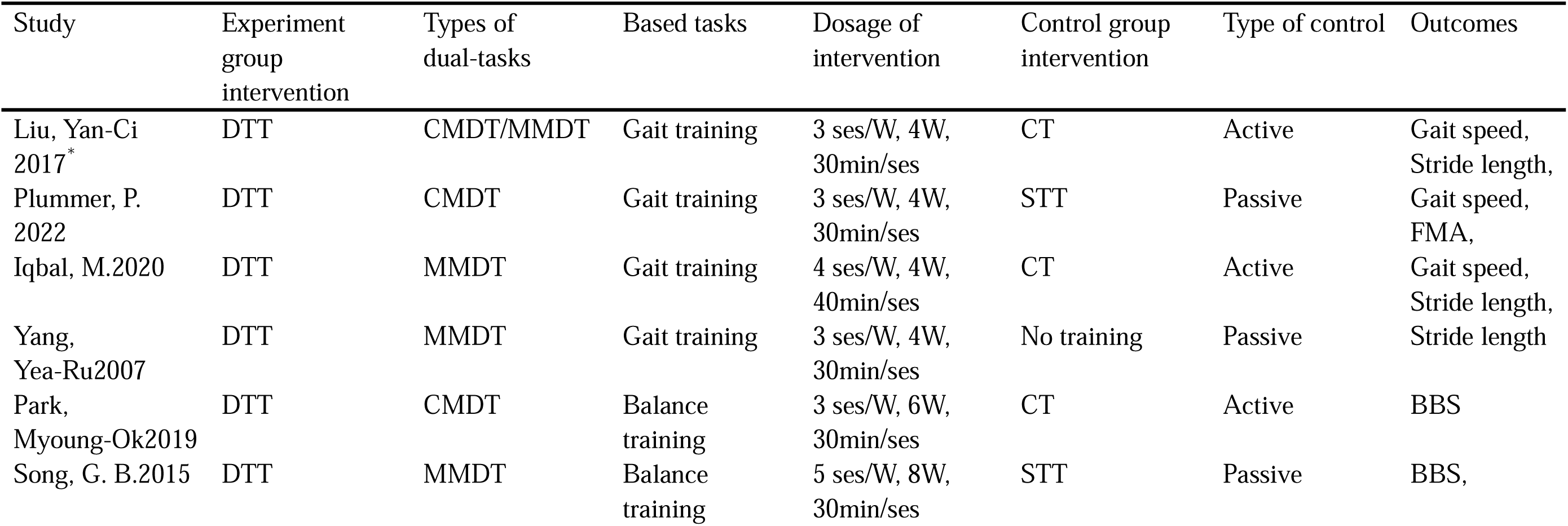
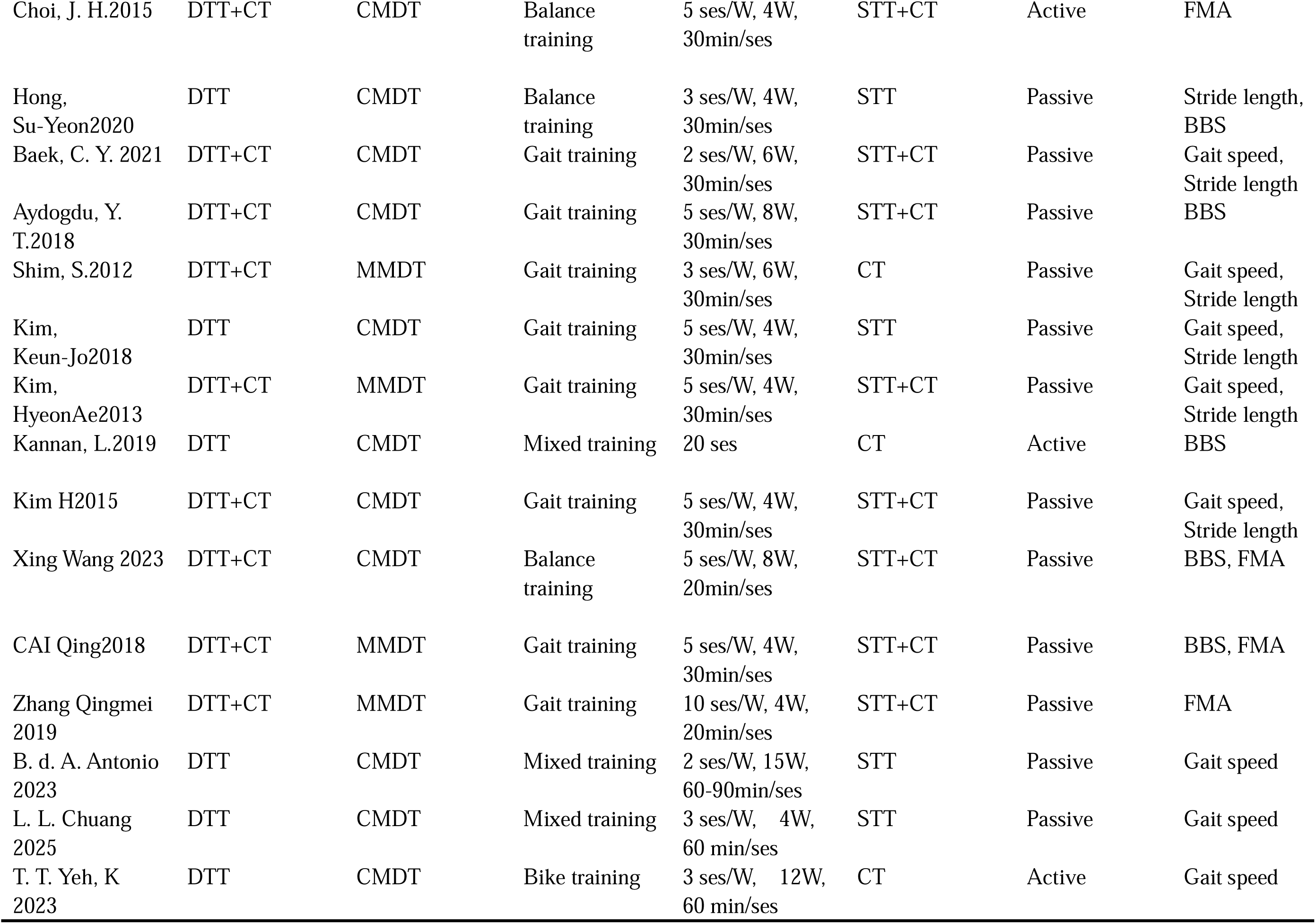
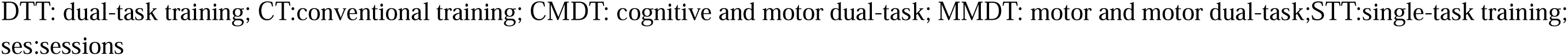
Characteristics of intervention.

| Study | Experiment group intervention | Types of dual-tasks | Based tasks | Dosage of intervention | Control group intervention | Type of control | Outcomes |
| --- | --- | --- | --- | --- | --- | --- | --- |
| Liu, Yan-Ci 2017* | DTT | CMDT/MMDT | Gait training | 3 ses/W, 4W, 30min/ses | CT | Active | Gait speed, Stride length, |
| Plummer, P. 2022 | DTT | CMDT | Gait training | 3 ses/W, 4W, 30min/ses | STT | Passive | Gait speed, FMA, |
| Iqbal, M.2020 | DTT | MMDT | Gait training | 4 ses/W, 4W, 40min/ses | CT | Active | Gait speed, Stride length, |
| Yang, Yea-Ru2007 | DTT | MMDT | Gait training | 3 ses/W, 4W, 30min/ses | No training | Passive | Stride length |
| Park, Myoung-Ok2019 | DTT | CMDT | Balance training | 3 ses/W, 6W, 30min/ses | CT | Active | BBS |
| Song, G. B.2015 | DTT | MMDT | Balance training | 5 ses/W, 8W, 30min/ses | STT | Passive | BBS, |
| Choi, J. H.2015 | DTT+CT | CMDT | Balance training | 5 ses/W, 4W, 30min/ses | STT+CT | Active | FMA |
| Hong, Su-Yeon2020 | DTT | CMDT | Balance training | 3 ses/W, 4W, 30min/ses | STT | Passive | Stride length, BBS |
| Baek, C. Y. 2021 | DTT+CT | CMDT | Gait training | 2 ses/W, 6W, 30min/ses | STT+CT | Passive | Gait speed, Stride length |
| Aydogdu, Y. T.2018 | DTT+CT | CMDT | Gait training | 5 ses/W, 8W, 30min/ses | STT+CT | Passive | BBS |
| Shim, S.2012 | DTT+CT | MMDT | Gait training | 3 ses/W, 6W, 30min/ses | CT | Passive | Gait speed, Stride length |
| Kim, Keun-Jo2018 | DTT | CMDT | Gait training | 5 ses/W, 4W, 30min/ses | STT | Passive | Gait speed, Stride length |
| Kim, HyeonAe2013 | DTT+CT | MMDT | Gait training | 5 ses/W, 4W, 30min/ses | STT+CT | Passive | Gait speed, Stride length |
| Kannan, L.2019 | DTT | CMDT | Mixed training | 20 ses | CT | Active | BBS |
| Kim H2015 | DTT+CT | CMDT | Gait training | 5 ses/W, 4W, 30min/ses | STT+CT | Passive | Gait speed, Stride length |
| Xing Wang 2023 | DTT+CT | CMDT | Balance training | 5 ses/W, 8W, 20min/ses | STT+CT | Passive | BBS, FMA |
| CAI Qing2018 | DTT+CT | MMDT | Gait training | 5 ses/W, 4W, 30min/ses | STT+CT | Passive | BBS, FMA |
| Zhang Qingmei 2019 | DTT+CT | MMDT | Gait training | 10 ses/W, 4W, 20min/ses | STT+CT | Passive | FMA |
| B. d. A. Antonio 2023 | DTT | CMDT | Mixed training | 2 ses/W, 15W, 60-90min/ses | STT | Passive | Gait speed |
| L. L. Chuang 2025 | DTT | CMDT | Mixed training | 3 ses/W, 4W, 60 min/ses | STT | Passive | Gait speed |
| T. T. Yeh, K 2023 | DTT | CMDT | Bike training | 3 ses/W, 12W, 60 min/ses | CT | Active | Gait speed |

DTT: dual-task training; CT:conventional training; CMDT: cognitive and motor dual-task; MMDT: motor and motor dual-task;STT:single-task training; ses:sessions

The final PEDro score ranged from five to eight points, with 80.96% of the included studies^15–17,26,28–41^ being high quality, scoring ≥ 6/10 , and 19.04% of the included studies^13,14,27,40^ being fair quality(Table 3)

**Table 3.** PEDro scale

| Study | Eligibility<br>criteria | Random<br>allocation | Concealed<br>allocation | Baseline comparability | Blind<br>subjects | Blind therapists | Blind assessors | Adequate follow-up | Intention<br>to-treat | Between-group comparisons | Point estimates<br>and variability | Total |
| --- | --- | --- | --- | --- | --- | --- | --- | --- | --- | --- | --- | --- |
| Liu,Yan-Ci2017 | 1 | 1 | 1 | 1 | 0 | 0 | 0 | 1 | 1 | 1 | 1 | 7/10 |
| Plummer,P.<br>2022 | 1 | 1 | 1 | 1 | 0 | 0 | 1 | 1 | 1 | 1 | 1 | 8/10 |
| Iqbal, M.<br>2020 | 1 | 1 | 1 | 1 | 0 | 0 | 0 | 1 | 1 | 1 | 1 | 7/10 |
| Yang,YeaRu<br>2007 | 1 | 1 | 1 | 1 | 0 | 0 | 1 | 1 | 0 | 1 | 1 | 7/10 |
| Park,<br>Myoung-Ok2019 | 1 | 1 | 1 | 1 | 0 | 0 | 1 | 1 | 1 | 1 | 1 | 8/10 |
| Song, G.B.<br>2015 | 1 | 1 | 0 | 1 | 0 | 0 | 0 | 1 | 1 | 1 | 1 | 6/10 |
| Choi, J.H.<br>2015 | 1 | 1 | 0 | 1 | 0 | 0 | 0 | 1 | 1 | 1 | 1 | 6/10 |
| Hong,<br>Su-Yeon2020 | 1 | 1 | 0 | 1 | 0 | 0 | 0 | 1 | 1 | 1 | 1 | 6/10 |
| Baek, C.Y. 2021 | 1 | 1 | 1 | 1 | 0 | 0 | 1 | 1 | 1 | 1 | 1 | 8/10 |
| Aydogdu, Y.<br>T.2018 | 1 | 1 | 0 | 1 | 0 | 0 | 0 | 0 | 1 | 1 | 1 | 5/10 |
| Shim, S.<br>2012 | 1 | 1 | 0 | 1 | 0 | 0 | 0 | 1 | 1 | 1 | 1 | 6/10 |
| Kim,K.J<br>2018 | 1 | 1 | 1 | 1 | 0 | 0 | 0 | 1 | 1 | 1 | 1 | 7/10 |
| Kim,HyeonAe<br>2013 | 1 | 1 | 0 | 1 | 0 | 0 | 0 | 1 | 1 | 1 | 1 | 6/10 |
| Kannan,L.<br>2019 | 1 | 1 | 0 | 1 | 0 | 0 | 0 | 1 | 1 | 1 | 1 | 6/10 |
| Kim H<br>2015 | 1 | 1 | 0 | 1 | 0 | 0 | 0 | 1 | 1 | 1 | 1 | 6/10 |
| Xing Wang 2023 | 1 | 1 | 0 | 1 | 0 | 0 | 0 | 0 | 1 | 1 | 1 | 5/10 |
| CAI Qing2018 | 1 | 1 | 0 | 1 | 0 | 0 | 1 | 0 | 1 | 1 | 1 | 6/10 |
| Zhang Qingmei<br>2019 | 1 | 1 | 0 | 1 | 0 | 0 | 1 | 0 | 1 | 1 | 1 | 6/10 |
| B. d. A. Antonio<br>2023 | 1 | 1 | 0 | 1 | 0 | 0 | 1 | 0 | 0 | 1 | 0 | 5/10 |
| L. L. Chuang<br>2025 | 1 | 1 | 0 | 0 | 0 | 0 | 1 | 1 | 0 | 1 | 0 | 5/10 |
| T. T. Yeh, K<br>2023 | 1 | 1 | 0 | 1 | 0 | 0 | 1 | 1 | 0 | 1 | 0 | 6/10 |

### Meta-analysis on gait performance

Twelve studies^13–17,31,33–36,38,41^ reported gait speed as the temporal gait parameter. One^36^ study included both MDT and CDT as intervention groups, allowing for two separate comparisons within the same study.

Therefore, a total of 13 comparisons were made between the experiment group and control group with considerable heterogeneity (I²=80%). Figure 2 shows a medium overall beneficial effect of dual-task training on gait speed compared to the control group (SMD=0.5, 95% CI = 0.05 - 0.95, p = 0.03). Due to limited comparisons or the presence of only a single category in some subgroups, subgroup analyses were not conducted for chronicity, basic motor training, or fall risk. Specifically, nearly all included studies recruited chronic patients; only two^14,15^ out of twelve studies did not involve gait training; and only five of the twelve studies reported measures of the BBS. In the subgroup analysis of walking ability, no significant differences were observed between the experimental and control groups for either home ambulators or community ambulators. Similarly, no significant subgroup differences were found based on the type of control. CDT did not demonstrate a significant advantage over conventional training in gait speed (SMD = 0.14, p=0.32). In contrast, MDT showed a significant improvement in gait speed compared to control groups, with a large effect size (SMD = 1.15) and a p-value of 0.01. This differential effect between MDT and CDT was also statistically supported by the meta-regression (p = 0.007), with a positive coefficient of 1.04 confirming the superior efficacy of MDT.

**Figure 2:**
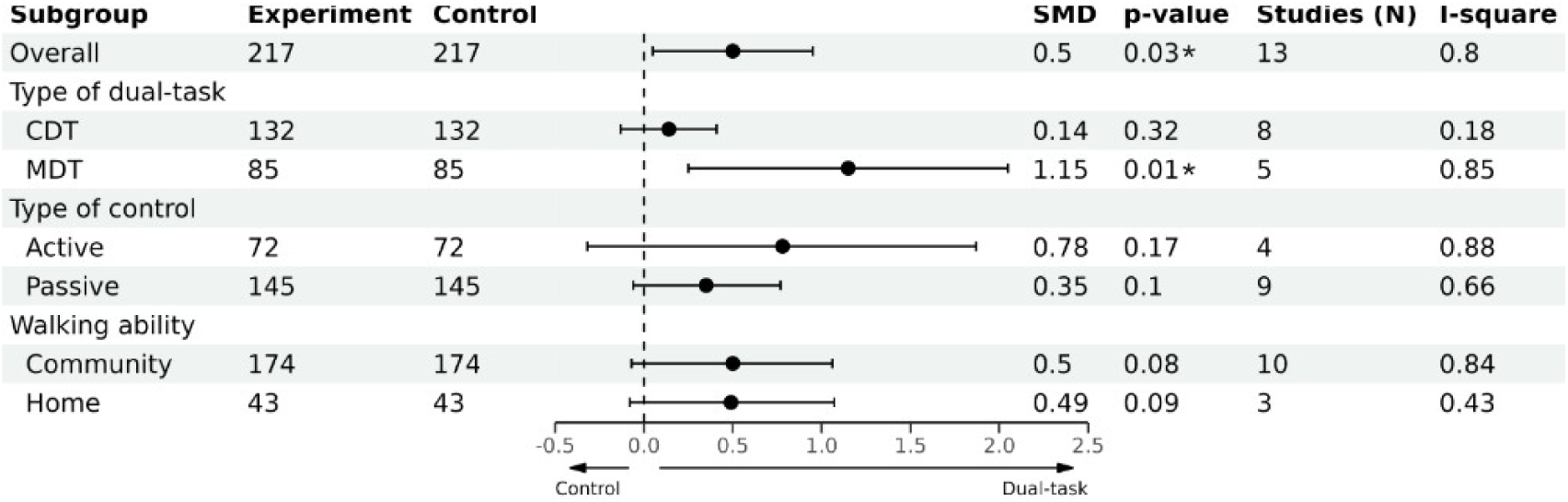
Gait performance--gait speed. SMD:Standardized Mean Difference. *Significant difference

As shown in Figure 3, eight studies^17,30,31,33–36,38,41^ were included, and nine comparisons were made to assess the effect of dual-task training on stride length. One study^33^ was excluded from the analysis due to substantial baseline discrepancies. The overall impact of dual-task training on stride length demonstrated a moderate effect with substantial heterogeneity (SMD = 0.5, 95% CI = 0.03–0.97, p=0.04, I²= 69%). In the subgroup analysis, MDT demonstrated a large effect size (SMD =0.89, p<0.01), whereas CDT showed no significant difference compared to the control group (SMD=0.01, p=0.97). Meta-regression analysis provided statistical support for dual-task type significantly moderating the treatment effect (p = 0.019), confirming that MDT demonstrated a statistically superior benefit, associated with a 0.90 larger effect size than CDT. In the subgroup analysis by control type, neither the active control group (p = 0.49) nor the passive control group (p = 0.06) showed statistically significant differences between experimental and control conditions. Likewise, in the walking ability subgroup analysis, no significant effects were observed for community ambulators (p = 0.06) or home ambulators (p = 0.30).

**Figure 3.**
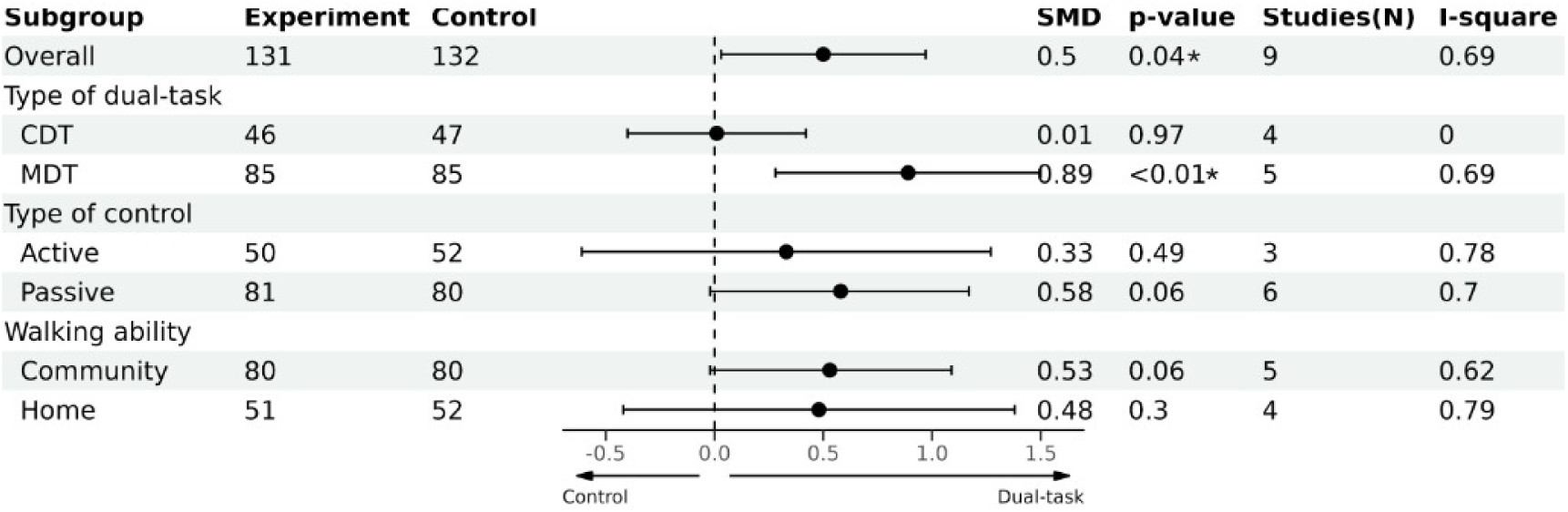
Gait performance—stride length. SMD:Standardized Mean Difference. *Significant difference

### Meta-analysis on balance control

Seven studies^27,28,30,32,37,39,40^ with significant heterogeneity (I² = 78%) were included and compared the effect of dual-task training on balance control (Figure 4). The combined results of the random-effect model were as follows: SMD=0.71, 95% CI = 0.14 - 1.28, p = 0.02, indicating a medium beneficial effect of dual-task training. In the subgroup analysis, patients with high fall risk showed greater improvement following dual-task training (SMD = 0.86, p < 0.01) compared to those with low fall risk (SMD = 0.54, p = 0.034), while no statistically significant effect was observed in patients with moderate fall risk (SMD = 0.84, p = 0.31). A significant training effect on BBS was observed in patients within 6 months post-stroke (SMD =1.82, p=0.034). This effect was not statistically significant in patients more than 6 months post-stroke (SMD = 0.29, p=0.08). Studies using CDT demonstrated a statistically significant improvement in balance control with a medium effect size (SMD=0.59, p<0.01). This training effect was not found in studies using MDT (SMD = 1.31, p = 0.35). Although the difference in effectiveness between CDT and MDT did not reach statistical significance in the meta-regression analysis (p = 0.064), the magnitude and direction of the effect sizes still suggest a preferential trend favoring the use of CDT for enhancing balance control.

**Figure 4:**
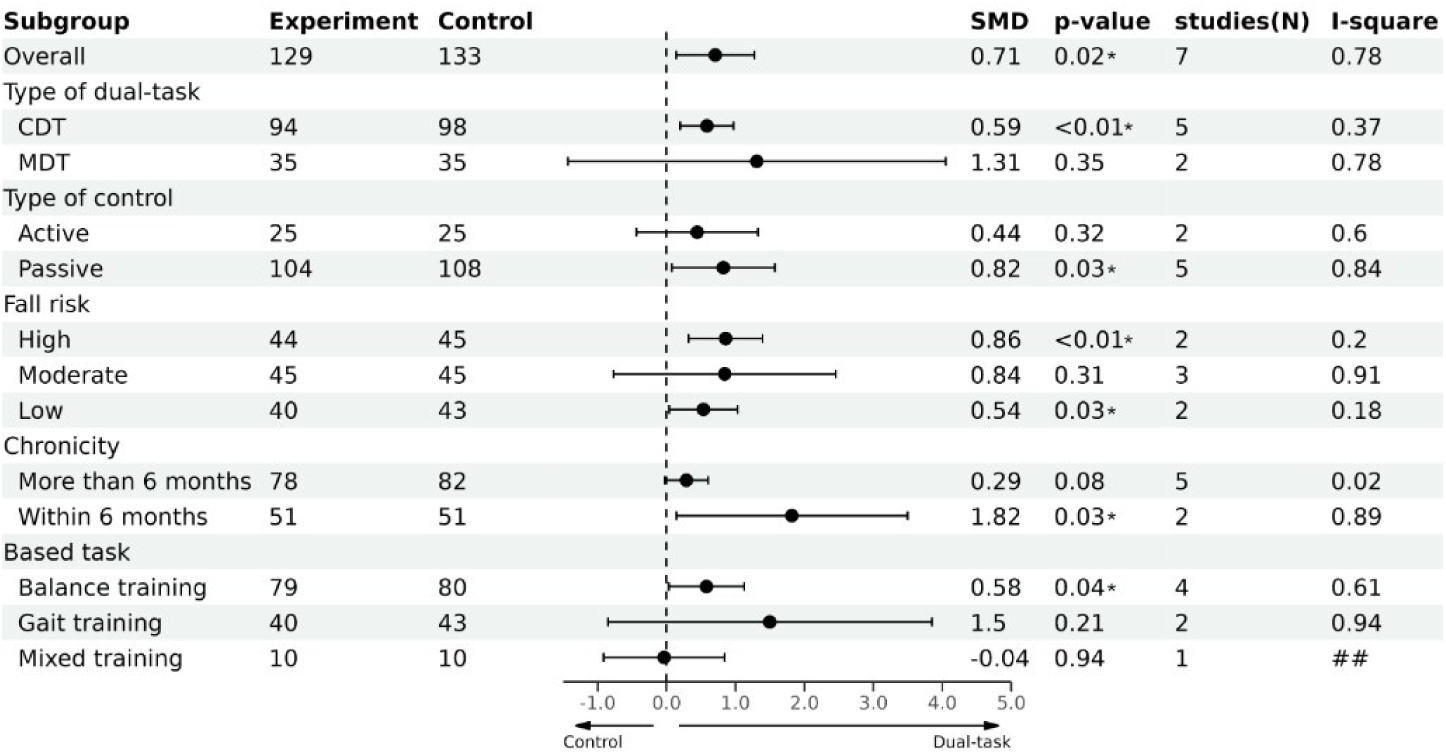
Berg balance scale. SMD:Standardized Mean Difference. *Significant difference

Compared to gait training and mixed training, balance training (SMD=0.579, p= 0.038) was the only one showing a significant difference between the experiment group and control group.

### Meta-analysis of motor function

As shown in Figure 5, five studies^16,26–29^ include FMA-LE as an outcome. No significant beneficial effect of dual-task training on motor function was found (SMD=0.42, 95% CI = −0.05–0.89, p = 0.08, I^2^ = 66%).

**Figure 5:**
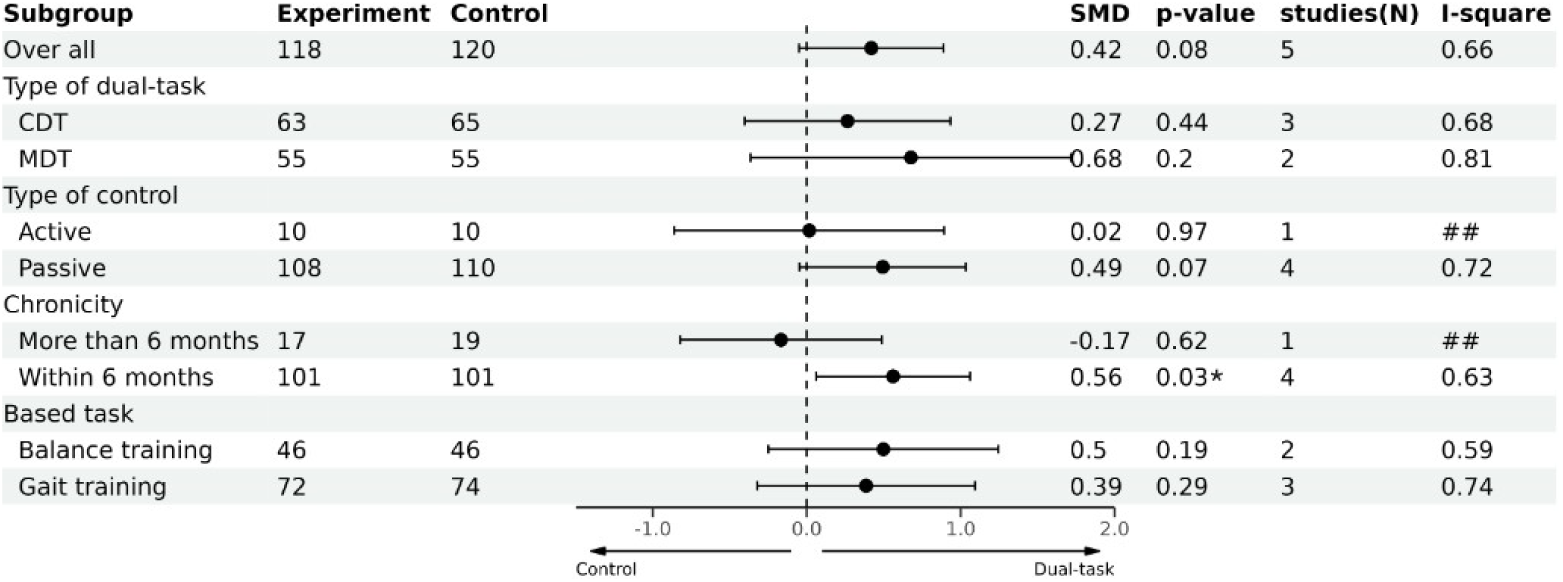
FMA-LE. SMD:Standardized Mean Difference. *Significant difference

Meta-regression analysis also did not find a statistically significant difference between CDT and MDT (p = 0.49), with a coefficient of 0.50. However, the subgroup analysis based on stroke chronicity again yielded a crucial finding for intervention timing: only patients in the subacute phase demonstrated a significant training effect (SMD= 0.56, p=0.03). No significant difference between the experimental group and control group was found in other subgroup analyses.

### Publication bias

As detailed in Table 4, Egger’s regression test indicated no significant evidence of publication bias across any of the main functional outcomes. The p-values for gait speed, stride length, BBS, and FMA-LE were 0.97, 0.39, 0.61, and 0.97, respectively, all well above the significance threshold of p < 0.05.

**Table 4.** Publication bias results of Egger’s test

| Outcome | Coef | Std.Err | 95% CI | t Value | P Value |
| --- | --- | --- | --- | --- | --- |
| Gait speed | -0.16 | 4.24 | -9.49 to 9.18 | -0.04 | 0.97 |
| Stride length | -2.9 | 3.14 | -10.33 to 4.53 | -0.92 | 0.39 |
| BBS | 1.79 | 3.3 | -6.68 to 10.26 | 0.54 | 0.61 |
| FMA-LE | 0.16 | 3.37 | -10.57 to 10.89 | 0.05 | 0.97 |

### Sensitivity analysis

The stability of the overall findings was confirmed through the sensitivity analysis using the leave-one-out method. Across all outcomes, none of the calculated pooled effect sizes fell outside the 95% confidence intervals, indicating that the overall findings of this meta-analysis are relatively stable and not influenced by any single study (Figure 6).

**Figure 6:**
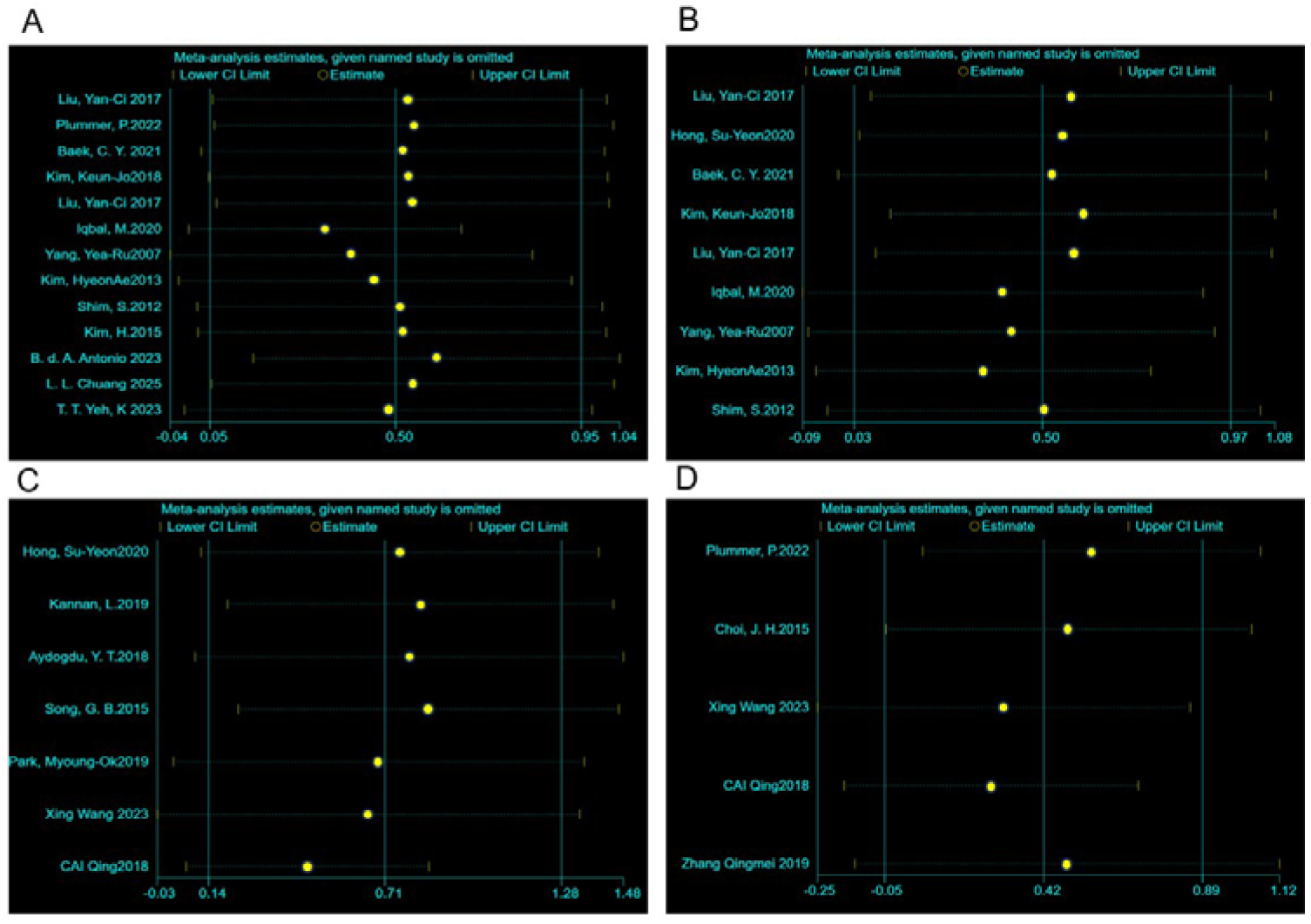
Sensitivity analysis of: A: gait speed; B: stride length; C: BBS; D: FMA_LE

## Discussion

The current meta-analysis systematically investigated the effects of dual-task training on physical recovery among stroke survivors at gait performance, balance control, and motor function, focusing on various factors that may influence the effectiveness of dual-task training, including the type of dual-task intervention, patient characteristics, and the underlying motor task. The primary finding confirmed that dual-task intervention offers a median effect over conventional treatment in improving both gait performance and balance control, though a consistent benefit was not observed for motor function. More importantly, the results provide clear guidance for a targeted prescription based on patients’ functional goals. MDT was found to be more effective than CDT for enhancing gait performance, whereas CDT seems to be better than MDT for improving balance control. Furthermore, patients’ characteristics may also contribute to the effectiveness of dual-task training. Patients within 6 months after stroke were more likely to gain improvement in balance control and physical function. Additionally, dual-task training had a greater effect on balance control in patients with a high risk of falls.

Three previous meta-analysis studies^6,7,42^ reported similar improvements on temporal gait parameters such as gait speed and cadence, which were consistent with our finding. There was mixed evidence in terms of spatial gait parameters such as stride length. Two of those studies ^6,42^ found the beneficial effect of dual-task training on spatial gait performance, which was also supported by our finding. The other one ^7^ reported no significant difference in stride length. The discrepancy may be attributed to the difference in inclusion criteria of the third study, as it included only studies employing CDT. However, none of those previous meta-analysis studies have specifically examined the differences in effectiveness between CDT and MDT.

Only a limited number of studies have explored this topic, but their findings have been inconsistent^10,11,43^. It has been speculated that different types of dual-task training might all positively impact walking performance but differently influence walking characteristics^10^. However, in the current study, we found significant effects only by MDT on both spatial and temporal gait parameters with a large effect size, whereas CDT did not demonstrate a significant influence on either parameter. Moreover, MDT was the only subgroup in the meta-analysis of gait performance to show a statistically significant difference among all subgroup analyses. These findings suggest that MDT may present significant training-specific effects, especially since the gait outcomes were single-task measures that did not involve simultaneous cognitive demands. One thing worth mentioning was that in the meta-analysis of gait performance, participants of nearly all included studies were chronic stroke survivors. Although no effect of CDT on gait performance was observed, it remains unclear whether CDT might improve gait performance in subacute stroke survivors.

There have been reports of mixed results of dual-task training on balance control in stroke survivors. One study didn’t observe the effect of dual-task training on balance control^3^, while other studies^44–46^ reported the superiority of dual-task training over conventional therapy. However, only one of the past three meta-analysis^7^ suggested the beneficial effect of dual-task training on balance control, not in the other two meta-analyses studies^6,42^. Our findings supported the overall beneficial effect of dual-task training on balance control in stroke survivors. Furthermore, we found that CDT provided greater improvements for balance control than MDT. The lack of observed effect of MDT on balance control may be attributed to the limitations of the BBS assessment, which does not adequately assess dynamic balance control as opposed to the Dynamic Gait Index (DGI)^47^. This is particularly relevant given that MDT is a more dynamic form of training. In subgroup analyses, we found that only balance training combined with a secondary task had a significant effect on balance control, whereas gait training and mixed training did not. This finding is reasonable, as it reflects the principle of task-specificity in training. The characteristics of patients could also influence the effectiveness of dual-task training, since stroke survivors with high fall risk showed the highest level of improvement in balance control, but participants with moderate fall risks didn’t show significant improvement. This result might be explained by the high heterogeneity of this subgroup (I² = 91%), compared to high fall risk (I² = 20%) and low fall risk (I² = 18%) subgroups. These findings collectively emphasize that for optimizing balance recovery, both the training modality (CDT) and the patient’s baseline fall risk profile must be carefully considered.

With more studies included in this meta-analysis, our results confirmed the finding of a previous meta-analysis^6^ that no superiority of dual-task training over conventional intervention on lower-extremity motor function. However, subgroup analysis provided a crucial prescriptive insight regarding the optimal timing of intervention. A significant benefit was found only in patients within 6 months post-stroke. This suggests that individuals in the subacute stage of recovery possess a higher level of neuroplasticity or residual function that makes them more responsive to the demands of dual-task interventions for improving lower-extremity motor function.

To the best of our knowledge, the current study is the first meta-analysis to perform subgroup analysis on the differential effects of CDT and MDT, as well as other factors that may influence the effectiveness of dual-task training. Such information can contribute to the goal of establishing targeted training protocols for stroke survivors, enabling clinicians to match the intervention modality to specific functional goals^10^. In addition, we included studies published in both English and Chinese. Only Chinese studies published in core journals are included to ensure the quality of the reviewed articles. Furthermore, the methodological integrity of our results was affirmed through sensitivity analysis. Despite moderate to substantial heterogeneities, the overall and subgroup findings for each outcome remained relatively stable when using the leave-one-out method, ensuring confidence in our interpretations.

This study also has several limitations. First, the beneficial effect of dual-task training should be interpreted with caution due to the heterogeneity of the reviewed trial studies. Second, the strength of evidence from subgroup analysis was limited by the small number of included studies. Additionally, the methodology of subgroup analysis did not allow for direct comparisons between categories. Therefore, the observed advantages of MDT over CDT in improving gait performance, and of CDT over MDT in enhancing balance, cannot be conclusively confirmed. More studies directly comparing these two types of dual-task training are needed to strengthen the evidence.

## Conclusion

The effectiveness of dual-task training in stroke rehabilitation is not uniform, suggesting the need to transition toward a precision rehabilitation approach that matches interventions to specific patient and task characteristics. Our review and meta-analysis investigated the effects of different types of dual-task training, along with other contributing factors such as the characteristics of stroke survivors. MDT appeared to produce greater benefits for gait performance, which was not observed with CDT. Conversely, CDT seemed to be more effective than MDT in enhancing balance function. The effectiveness of dual-task training also varied based on patient characteristics; Subacute stroke survivors in the subacute phase were more likely to experience improvements in balance control and motor function compared to those at the chronic stage.

Stroke survivors with a high risk of falls were identified as optimal subgroups likely to experience greater gains in balance control and motor function. These findings offer crucial prescriptive insights for clinicians, guiding the selection of the dual-task modality and the timing of intervention based on the individual stroke survivor’s baseline function profile. However, the high heterogeneity among studies and the lack of direct comparisons between CDT and MDT limit the strength of these conclusions. Future research should aim to further clarify the specific benefits of different types of dual-task training and identify the most suitable populations to maximize therapeutic outcomes.

## Data Availability

All data produced in the present study are available upon reasonable request to the authors.

## Acknowledgments

The author(s) received no financial support for the research and authorship.

